# Effect of Acute Kidney Injury on In-hospital Mortality in Non-critical Medical Patients in a Sub-Saharan African Country

**DOI:** 10.1101/2023.09.20.23295826

**Authors:** Nahom Desalegn Mekonnen, Tigist Workneh Leulseged, Nebiat Adane Mera, Helen Surafeal Berhe, Anteneh Abera Beyene, Kidus Haile Yemaneberhan, Buure Ayderuss Hassen, Feven Negasi Abriha, Lidiya Zenebe Getachew, Birukti Gebreyohannes Habtezgi

**Affiliations:** Department of Internal Medicine, St. Paul’s Hospital Millennium Medical College, Addis Ababa, Ethiopia; Medical Research Lounge (Research Consultancy and Training Center), Addis Ababa, Ethiopia; Department of Internal medicine, Yekatit 12 Hospital Medical College, Addis Ababa, Ethiopia; Afya Clinic, Kansas City, Missouri; MyungSung Medical College comprehensive specialized hospital Addis Ababa, Ethiopia; Hayat medical college, Addis Ababa, Ethiopia; Jimma University school of medicine, Jimma, Ethiopia; Eka Kotebe General Hospital, Addis Ababa, Ethiopia

**Keywords:** Acute kidney Injury, non-critical setting, retrospective cohort, log binomial regression, Ethiopia

## Abstract

**Background:** AKI is a major global public health problem that affects millions of people each year and has been linked to poor prognosis in critically ill patients. As being a common complication in hospitalized patients, understanding its effect on non-critical patients is equally crucial, but there is a paucity of knowledge in this area, particularly in Africa. Therefore, the aim of this study was to assess the effect of AKI on in-hospital morality in non-critical medical patients admitted to a large tertiary hospital in Ethiopia.

**Methods:** A retrospective cohort study of 319 non-critical medical patients (113 with AKI and 206 without AKI) admitted between July 2019 and January 2022 was conducted. The in-hospital mortality rate was estimated using incidence density with a 95% CI. The two groups’ comparability was assessed using chi-square and Fisher’s exact tests. The effect of AKI on in-hospital mortality was analyzed using a log binomial regression model with a p-value of ≤ 0.05 determining a significant effect, and the effect was measured using adjusted relative risk (ARR) and its 95% CI.

**Results:** The in-hospital mortality rate was 6.8 per 1000 person-days of observation (95% CI=4.9-9.4). AKI did not show a significant effect on in-hospital mortality (ARR = 0.72, 95% CI=0.30-1.71, p=0.450). On the other hand, sepsis was found to be a significant predictor, with over a threefold increase in risk of in-hospital mortality (ARR=3.47, 95% CI=1.60-7.52, p=0.002).

**Conclusion:** With early detection and proper management, non-critical patients with AKI can have a similar prognosis as those without AKI, unlike the critical setting. However, sepsis was found to be a significant predictor of in-hospital mortality implying the need to pay special attention to the management of these cases.

## INTRODUCTION

Acute kidney injury (AKI) is a sudden decrease in kidney function that occurs over hours or days and can be caused by structural or functional damage to the kidneys (1). AKI is a major global public health problem affecting over 13 million people every year (2, 3). It is a frequent complication among patients with other medical conditions, whether it occurs in the community or the hospital setting (4-9).

In an ideal hospital setting most patients with AKI make a full recovery. However, in resource-constrained settings, where early diagnosis and treatment are compromised by limited resources and patients often delay seeking care, AKI can lead to prolonged hospitalization, complications, and increased mortality. In particular, critical patients with AKI have a high mortality rate, up to 23.9%. It is reported that patients with AKI have up to 10 times increased risk of death as compared to patients without AKI (3, 10-16).

It is reported that Sub-Saharan Africa has a higher incidence of AKI than the rest of the world. Because of the underdeveloped health-care infrastructure, which causes gaps in the management of these cases, disease progression and prognosis are also worse, resulting in higher rates of complication and death. According to studies, patients with AKI in these settings have a death rate of up to 36.9%, which is five times greater than the death rate in patients without AKI (7, 10, 15, 17-22).

However, because these studies were predominantly carried out in developed contexts, their scope and generalizability are limited, and their findings may not be applicable to all settings in Sub-Saharan Africa. Aside from the scarcity of evidence in Africa and the epidemiological variance in disease patterns and outcomes across different geographic locations, the majority of previous studies were conducted on critical patients who required ICU care. However, AKI is a also common consequence in non-critical patients, and hence it is crucial to investigate the impact of this disease on treatment outcomes in these group of patients. Therefore, the aim of this study was to assess the effect of AKI on in-hospital morality in non-critical medical patients admitted to St. Paul’s Hospital Millennium Medical College between July 2019 and January 2022.

## METHODS

### Study Setting and Design

An institution-based retrospective cohort study was conducted among non-critical medical patients admitted to the medical ward of St. Paul’s Hospital Millennium Medical College (SPHMMC) between July 2019 and January 2022, comparing patients with AKI (exposed group) to those without AKI (non-exposed group). SPHMMC is one of the largest hospitals in Ethiopia. It is a center that specializes in renal care and treatment with the only renal transplant facility in the country since 2015. The center has a well-equipped center for dialysis that gives special attention to patients with acute kidney injury. The unit has 8 nephrologists, 4 fellows and over 70 nurses providing comprehensive inpatient and outpatient care. The conduct and findings of the entire research is reported in accordance with the STROBE checklist for cohort studies.

### Population and Sample Size

The study included all eligible non-critical medical patients who were admitted to the hospital between July 2019 and January 2022. Patients were considered eligible if they did not have an underlying chronic renal condition at the time of admission, completed their follow-up at the hospital, were not transferred from or admitted to a critical ward or intensive care unit at any point during their stay in the hospital, and had a complete medical record on the major exposures and outcome variables. Accordingly, a total of 319 eligible cases were identified and enrolled in the study, 113 patients with AKI (45 Stage 1, 16 Stage 2, and 52 Stage 3) and 206 patients without AKI.

**Figure 1:**
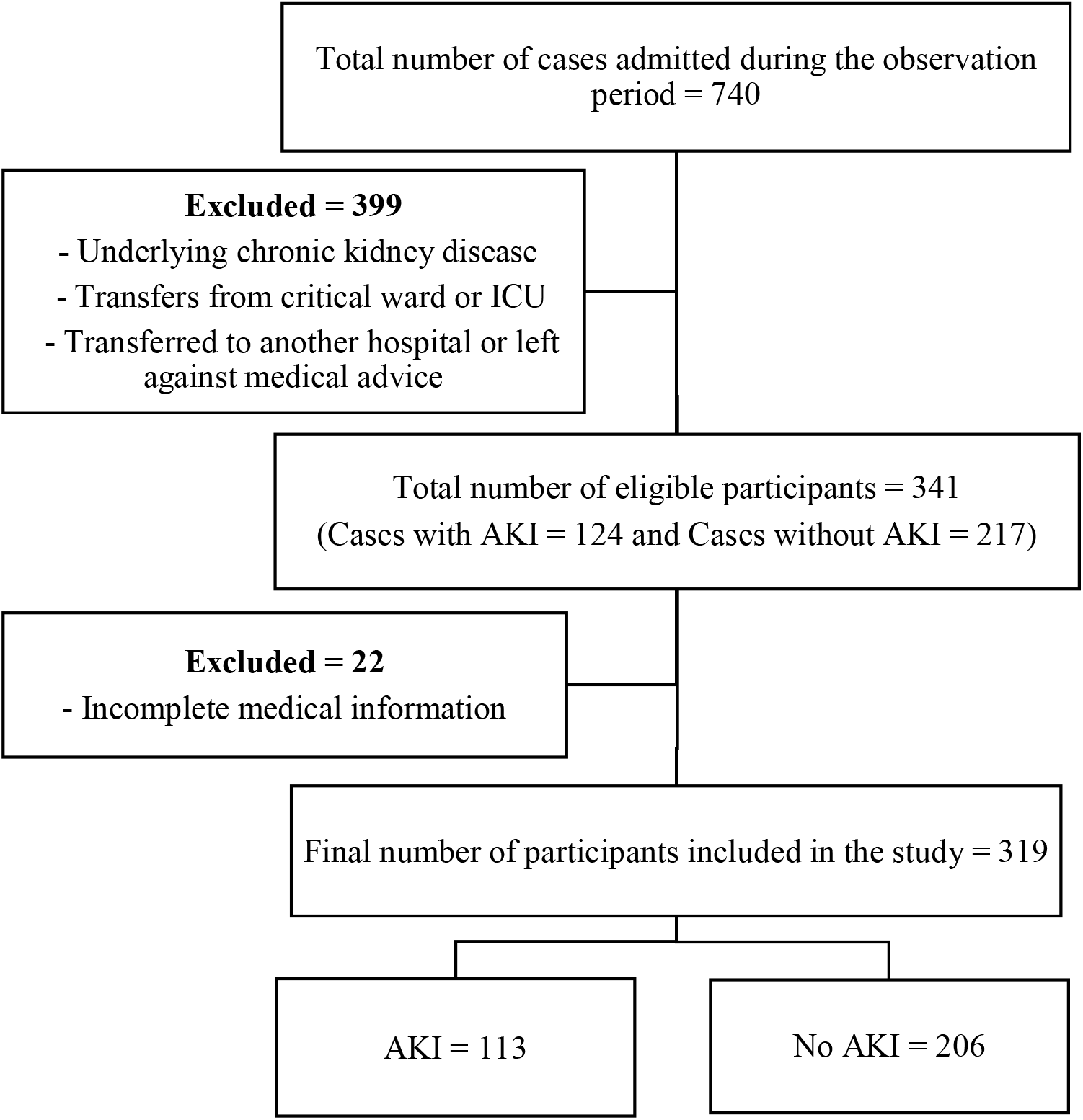
Flow chart showing the recruitment of study participants in to the study, July 2019 to January 2022, Ethiopia.

### Operational Definition

#### Acute Kidney Injury (AKI)

Diagnosed based on Kidney Disease Improving Global Outcomes criteria (KDIGO) definition, is defined as an increase in serum creatinine by ≥ 0.3mg/dl within 48 hours or increase in serum creatinine to ≥ 1.5 times baseline, which is known or presumed to have occurred within the previous 7 days (23).

- **Stage 1:** an increase in serum creatinine to 1.5 to 1.9 times baseline, or increase in serum creatinine by ≥0.3 mg/dL, or reduction in urine output to <0.5 mL/kg/hour for 6 to 12 hours.
- **Stage 2:** an increase in serum creatinine to 2.0 to 2.9 times baseline, or reduction in urine output to <0.5 mL/kg/hour for ≥12 hours
- **Stage 3**: an increase in serum creatinine to 3.0 times baseline, or increase in serum creatinine to ≥4.0 mg/dL or reduction in urine output to <0.3 mL/kg/hour for ≥24 hours, or anuria for ≥12 hours, or the initiation of kidney replacement therapy

### Data Collection Procedures and Quality Assurance

Data on the exposures and the study outcome were extracted from the patients’ medical charts using a pre-tested data abstraction form. Three General Practitioners collected data under the supervision of a senior Internal Medicine resident. To ensure data quality, data collectors and the supervisor were trained on how to use the tool, data inconsistency was identified by running frequencies and cross tabulations and managed by cross checking with the charts, and additional data cleaning and management was performed for variables with numeric errors and missing values. STATA software version 17.0 (College Station, TX) was used for all data management and analysis.

### Statistical Analysis

The data was summarized and presented using a frequency table with percentage. The follow-up duration of the patients was summarized using median with interquartile range, due to the skewed nature of the data (p-value of <0.0001 for Kolmogorov-Smirnov and Shapiro Wilk tests of normality). To calculate the incidence of in-hospital mortality rate, incidence density with 95% CI was calculated with the total person time observation measured in days.

The underlying socio-demographic and clinical characteristics of the two groups (AKI Vs Without AKI) were compared using a Chi-square test after testing the assumptions of the test. For variables with failed assumptions, Fisher’s exact test was used instead. For both tests, a p-value of ≤ 0.05 indicates the presence of a statistically significant difference between the groups.

To identify the effect of AKI on in-hospital mortality, a multivariable log binomial regression model was fitted after adjusting for variables that passed the statistical selection criteria of univariate analysis at 25% level of significance and that are found to be scientifically relevant. On the multivariable model, a significant effect was determined at a p-value of ≤ 0.05 and the effect was measured using adjusted relative risk (ARR) and its 95% CI.

## RESULTS

### Socio-demographic and Clinical Characteristics

More than half of the patients were under the age of 50, with the majority being younger than 30 years (22.6%) and between 30 and 39 years (18.2%), and 173 (54.2%7) were males. Hypertension and type 2 diabetes mellitus (T2DM) were the most frequently reported comorbid illnesses in 138 (43.3%) and 55 (17.2%) individuals, respectively. Pneumonia was the most common admission diagnosis in 112 (35.15%), followed by heart failure in 76 (23.8%), hypercoagulable states (DVT/PE) in 46 (14.4%), and sepsis in 40 (12.5%).

When the underlying socio-demographic and clinical characteristics of patients without AKI and those with AKI were compared, it was found that a significant majority of patients with AKI were hypertensive (36.4% vs 55.8%, p-value=0.001) and had sepsis on admission (9.2% vs 18.6%, p=0.016). On the other hand, pneumonia (39.8% vs 26.5%, p=0.018), CNS infection (8.7% vs 0, p=0.001), stroke (12.6% vs 4.4%, p=0.018), DVT/PE (17.5% vs 8.8%, p=0.036), and hematologic disorders (7.3% vs 0.9%, p=0.012) were seen in a significantly smaller proportion of AKI patients. Otherwise, there was no significant difference between the two groups in terms of age, sex, or other clinical factors. **(Table 1)**

**Table 1:** Socio-demographic and clinical characteristics of non-critical medical patients and comparison based on the diagnosis of AKI, July 2019 to January 2022, Ethiopia (n=319)

| Variable |  | Total (%) | AKI Diagnosis |  | p-value |
| --- | --- | --- | --- | --- | --- |
|  |  |  | No (%)<br>(n=206) | Yes (%)<br>(n=113) |  |
| Age group (in years) | <30 | 72 (22.6) | 49 (23.8) | 23 (20.4) | 0.829 |
|  | 30-39 | 58 (18.2) | 36 (17.5) | 22 (19.5) |  |
|  | 40-49 | 48 (15.0) | 34 (16.5) | 14 (12.4) |  |
|  | 50-59 | 50 (15.7) | 30 (14.6) | 20 (17.7) |  |
|  | 60-69 | 51 (16.0) | 31 (15.0) | 20 (17.7) |  |
|  | ≥70 | 40 (12.5) | 26 (12.6) | 14 (12.7) |  |
| Sex | Male | 173 (54.2) | 112 (54.4) | 61 (54.0) | 0.947 |
|  | Female | 146 (45.8) | 94 (45.6) | 52 (46.0) |  |
| Hypertension | No | 181 (56.7) | 131 (63.3) | 50 (44.2) | 0.001* |
|  | Yes | 138 (43.3) | 75 (36.4) | 63 (55.8) |  |
| Type 2 Diabetes | No | 264 (82.8) | 174 (84.5) | 90 (79.6) | 0.276 |
| **Mellitus** | **Yes** | 55 (17.2) | 32 (15.5) | 23 (20.4) |  |
| **HIV** | **No** | 304 (95.3) | 193 (93.7) | 111 (98.2) | 0.067 |
|  | **Yes** | 15 (1.8) | 13 (6.3) | 2 (1.8) |  |
| **Malignancy** | **No** | 307 (96.2) | 197 (95.6) | 110 (97.3) | 0.442 |
|  | **Yes** | 12 (3.8) | 9 (4.4) | 3 (2.7) |  |
| **Sepsis** | **No** | 279 (87.5) | 187 (90.8) | 92 (81.4) | **0.016\*** |
|  | **Yes** | 40 (12.5) | 19 (9.2) | 21 (18.6) |  |
| **Heart failure** | **No** | 243 (76.2) | 157 (76.2) | 86 (76.1) | 0.983 |
|  | **Yes** | 76 (23.8) | 49 (23.8) | 27 (23.9) |  |
| **Pneumonia** | **No** | 207 (64.9) | 124 (60.2) | 83 (73.5) | **0.018\*** |
|  | **Yes** | 112 (35.1) | 82 (39.8) | 30 (26.5) |  |
| **TB** | **No** | 293 (91.8) | 186 (90.3) | 107 (94.7) | 0.170 |
|  | **Yes** | 26 (8.2) | 20 (9.7) | 6 (5.3) |  |
| **COPD** | **No** | 309 (96.9) | 198 (96.1) | 111 (98.2) | 0.504 |
|  | **Yes** | 10 (3.1) | 8 (3.9) | 2 (1.8) |  |
| **Hepatitis** | **No** | 295 (92.5) | 193 (93.7) | 102 (90.3) | 0.268 |
|  | **Yes** | 24 (7.5) | 13 (6.3) | 11 (9.7) |  |
| **GI infection** | **No** | 306 (95.9) | 197 (95.6) | 109 (96.5) | 0.999 |
|  | **Yes** | 13 (4.1) | 9 (4.4) | 4 (3.5) |  |
| **Upper GI Bleeding** | **No** | 308 (96.6) | 198 (96.1) | 110 (97.3) | 0.752 |
|  | **Yes** | 11 (3.4) | 8 (3.9) | 3 (2.7) |  |
| **Gastric/duodenal ulcer** | **No** | 312 (97.8) | 201 (97.6) | 111 (98.2) | 0.999 |
|  | **Yes** | 7 (2.2) | 5 (2.4) | 2 (1.8) |  |
| **CNS infection** | **No** | 301 (94.4) | 188 (91.3) | 113 (100.0) | **0.001\*** |
|  | **Yes** | 18 (5.6) | 18 (8.7) | 0 |  |
| **Stroke** | **No** | 288 (90.3) | 180 (87.4) | 108 (95.6) | **0.018\*** |
|  | **Yes** | 31 (9.7) | 26 (12.6) | 5 (4.4) |  |
| **DVT/PE** | **No** | 273 (85.6) | 170 (82.5) | 103 (91.2) | **0.036\*** |
|  | **Yes** | 46 (14.4) | 36 (17.5) | 10 (8.8) |  |
| **Hematologic disease** | **No** | 303 (95.0) | 191 (92.7) | 112 (99.1) | **0.012\*** |
|  | **Yes** | 16 (5.0) | 15 (7.3) | 1 (0.9) |  |

N.B., \* = Statistically significant

### Medication history and Laboratory Parameters

Over a third of the patients were taking one or a combination of the following medications: cephalosporin (77.7%), proton pump inhibitors (PPIs) (60.8%), anticoagulants (54.2%), diuretics (39.8%), and vancomycin (38.6%). Low white blood cell count (WBC), hemoglobin, Sodium, potassium, and chloride levels were found in 31 (9.7%), 151 (47.3%), 112 (35.1%), 51 (16.0%), and 104 (32.6%) of the patients, respectively. Furthermore, elevated levels of WBC, hemoglobin, sodium, potassium, and chloride were detected in 86 (27.0%), 38 (11.9%), 10 (3.1%), 29 (9.1%), and 104 (32.6%) patients, respectively.

The medication history and the baseline laboratory parameters between the cohorts revealed that a significantly larger proportion of patients with AKI were on diuretics (25.7% Vs 65.5%, p<0.0001) and PPIs (53.4% Vs 74.3%, p<0.0001) than those without AKI. Furthermore, a significantly larger proportion of these AKI patients had abnormal baseline laboratory parameters, including anemia (40.0% Vs 59.3%, p=0.003), hyponatremia (28.2% Vs 47.8%, p=0.006), hyperkalemia (3.9% Vs 18.6%, 0.0001), hypochloremia (27.7% Vs 41.6%, p=0.002), and hyperchloremia (5.8% Vs 11.5%, p=0.002). **(Table 2)**

**Table 2:** Medication history and laboratory parameters of non-critical medical patients and comparison based on the diagnosis of AKI, July 2019 to January 2022, Ethiopia (n=319)

| Variable |  | Total (%) | AKI Dx |  | p-value |
| --- | --- | --- | --- | --- | --- |
|  |  |  | No (%)<br>(n=206) | Yes (%)<br>(n=113) |  |
| Cephalosporin | No | 71 (22.3) | 51 (24.8) | 20 (17.7) | 0.147 |
|  | Yes | 248 (77.7) | 155 (75.3) | 93 (82.3) |  |
| Penicillin | No | 300 (94.0) | 193 (93.7) | 107 (94.7) | 0.718 |
|  | Yes | 19 (6.0) | 13 (6.3) | 6 (5.3) |  |
| Macrolides | No | 273 (85.6) | 173 (84.0) | 100 (88.5) | 0.272 |
|  | Yes | 46 (14.4) | 33 (16.0) | 13 (11.5) |  |
| Quinolones | No | 307 (96.2) | 200 (97.1) | 107 (94.7) | 0.282 |
|  | Yes | 12 (3.8) | 6 (2.9) | 6 (5.3) |  |
| Vancomycin | No | 196 (61.4) | 131 (63.6) | 65 (57.7) | 0.287 |
|  | Yes | 123 (38.6) | 75 (36.4) | 48 (42.5) |  |
| Anti-TB | No | 289 (90.6) | 184 (89.3) | 105 (92.9) | 0.292 |
|  | Yes | 30 (9.4) | 22 (10.7) | 8 (7.1) |  |
| Diuretics | No | 192 (60.2) | 153 (74.3) | 39 (34.5) | <0.0001* |
|  | Yes | 127 (39.8) | 53 (25.7) | 74 (65.5) |  |
| ACEI/ARBs | No | 253 (79.3) | 170 (82.5) | 83 (73.5) | 0.056 |
|  | Yes | 66 (20.7) | 36 (17.5) | 30 (26.5) |  |
| PPIs | No | 125 (39.2) | 96 (46.6) | 29 (25.7) | <0.0001* |
|  | Yes | 194 (60.8) | 110 (53.4) | 84 (74.3) |  |
| Anticoagulants | No | 146 (45.8) | 91 (44.2) | 55 (48.7) | 0.441 |
|  | Yes | 173 (54.2) | 115 (55.8) | 58 (51.3) |  |
| Steroids | No | 220 (69.0) | 148 (71.8) | 72 (63.7) | 0.133 |
|  | Yes | 99 (31.0) | 58 (28.2) | 41 (36.3) |  |
| WBC | Normal | 202 (63.3) | 133 (64.6) | 69 (61.1) | 0.228 |
|  | Low | 31 (9.7) | 23 (11.2) | 8 (7.1) |  |
|  | High | 86 (27.0) | 50 (24.3) | 36 (31.9) |  |
| Hemoglobin | Normal | 130 (40.8) | 98 (47.6) | 32 (28.3) | 0.003* |
|  | Low | 151 (47.3) | 84 (40.8) | 67 (59.3) |  |
|  | **High** | 38 (11.9) | 24 (11.7) | 14 (12.4) |  |
| **Sodium** | **Normal** | 197 (61.8) | 141 (68.4) | 56 (49.6) | **0.006\*** |
|  | **Low** | 112 (35.1) | 58 (28.2) | 54 (47.8) |  |
|  | **High** | 10 (3.1) | 7 (3.4) | 3 (2.7) |  |
| **Potassium** | **Normal** | 239 (74.9) | 166 (80.6) | 73 (64.6) | **<0.0001\*** |
|  | **Low** | 51 (16.0) | 32 (15.5) | 19 (16.8) |  |
|  | **High** | 29 (9.1) | 8 (3.9) | 21 (18.6) |  |
| **Chloride** | **Normal** | 190 (59.6) | 137 (66.5) | 53 (46.9) | **0.002\*** |
|  | **Low** | 104 (32.6) | 57 (27.7) | 47 (41.6) |  |
|  | **High** | 25 (7.8) | 12 (5.8) | 13 (11.5) |  |

N.B., \* = Statistically significant

### Incidence of In-hospital Mortality

The 319 patients were followed for a median duration of 13.0 days (IQR, 8.0-20.0). The overall in-hospital mortality rate (MR) was 6.8 per 1000 person-days (PD) of observation (95% CI=4.9 - 9.4). Based on the sub-group analysis, the MR in those without AKI was 6.8 per 1000 PD (95% CI=4.4-10.4) and among those with AKI it was 6.8 per 1000 PD (95% CI=4.0-11.4).

### Effect of AKI on In-hospital mortality

To assess the effect of AKI on in-hospital mortality, a multivariable Log binomial Regression model was fitted after adjusting for age group, sex, hypertension, T2DM, sepsis, heart failure, pneumonia, DVT/PE, cephalosporin, vancomycin, diuretics, ACEIs/ARBs, PPIs, and steroids. Accordingly, AKI was not found to be a significant predictor of in-hospital mortality **(ARR = 0**.**72, 95% CI=0**.**30-1**.**71, p=0**.**450)**.

From the other exposures, sepsis was found to be the only significant predictor for in-hospital mortality, where patients with sepsis had a 3.47 times increased risk of in-hospital mortality than those with no sepsis **(ARR=3**.**47, 95% CI=1**.**60-7**.**52, p=0**.**002)**. (**Table 3**)

**Table 3:**
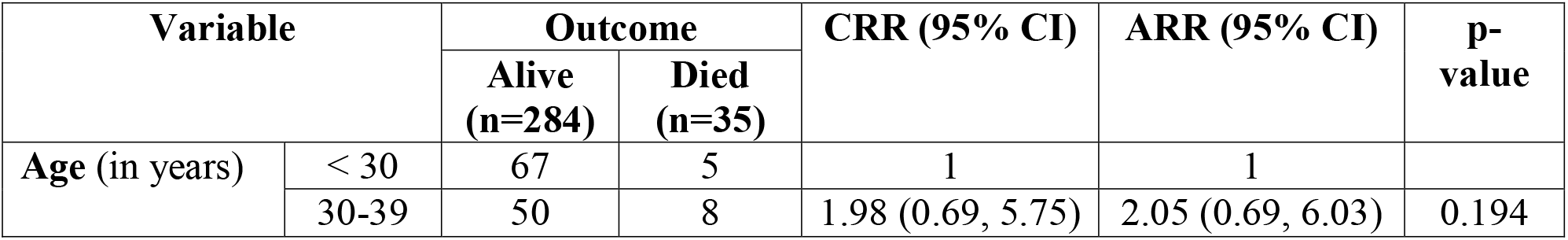
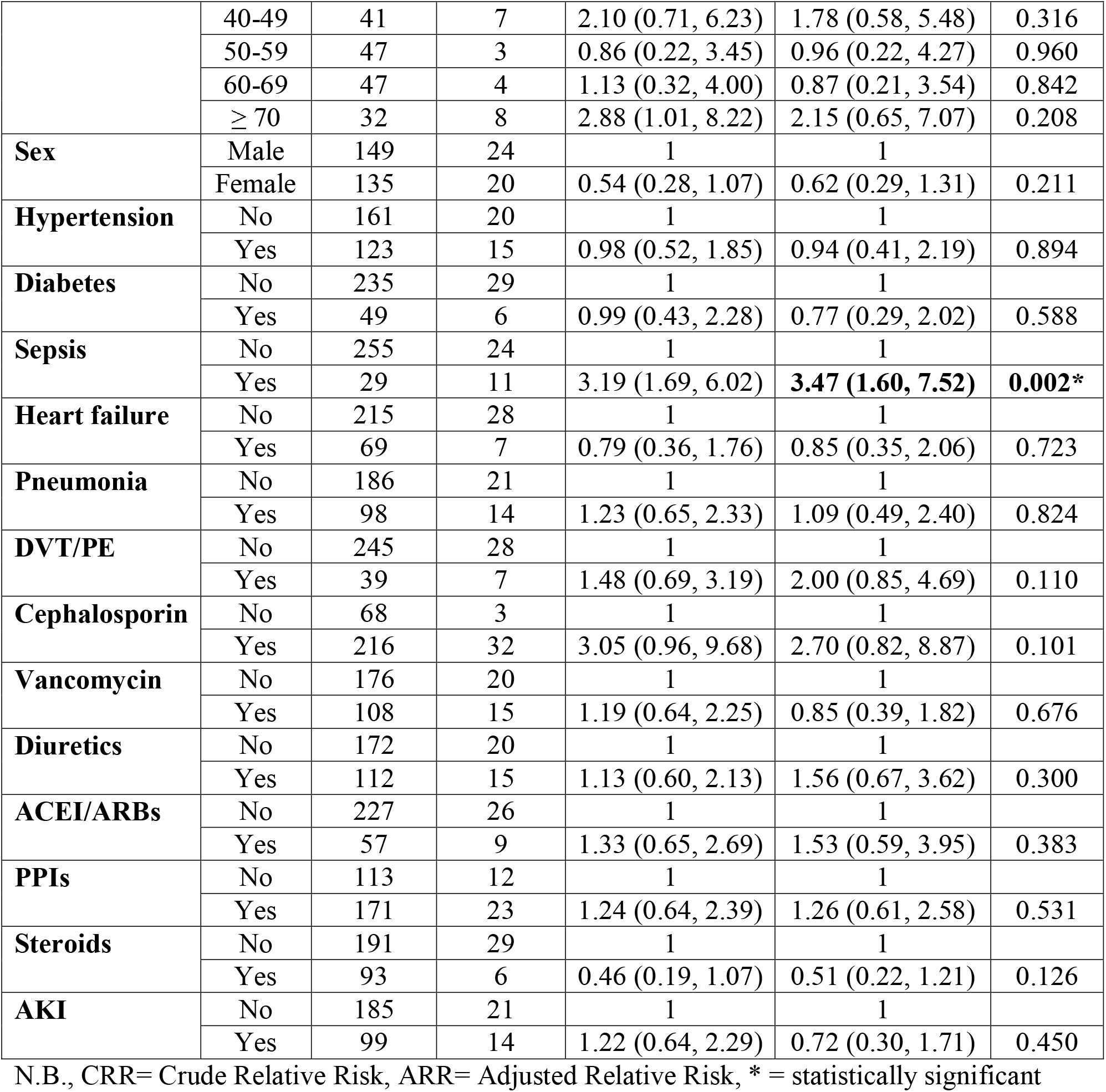
Determinants of in-hospital mortality among non-critical medical patients, July 2019 to January 2022, Ethiopia (n=319)

## DISCUSSION

The current study was conducted on 319 non-critical medical patients, 113 patients with AKI and 206 patients without AKI, admitted to one of Ethiopia’s largest tertiary hospitals in order to determine the effect of AKI on in-hospital mortality. The majority of the patients were males (54.2%) and under the age of 50 (53.3%). Comparison within the cohort showed that, the significant majority of AKI patients were hypertensive, had sepsis at the time of admission, were on diuretics and PPIs, and had abnormal baseline laboratory values including anemia, hyponatremia, hyperkalemia, hypochloremia, and hyperchloremia. Pneumonia, CNS infection,stroke, DVT/PE, and hematologic abnormalities, on the other hand, were seen in a much lower proportion of patients with AKI.

The in-hospital mortality rate was determined to be 6.8 per 1000 PD and the incidence rate remained the same in sub-group analysis, demonstrating that there was no significant difference in the mortality rate between patients with and without AKI. Further regression analysis to examine AKI’s predictive effect on in-hospital mortality revealed that AKI was not a significant predictor of in-hospital mortality. In contrast, evidence in critical care settings in both developed and developing countries shows that having AKI worsens disease progression and increases mortality by up to tenfold (10-16, 17-22). Hence, the findings of this study implies that the detrimental effects of AKI may be less severe in non-critical patients. However, it is important to note that the underlying medical conditions of patients without AKI in our study were more severe, which may have confounded the results. Furthermore, the hospital’s setup for AKI management could have contributed to earlier identification and better management of AKI cases, improving their outcome.

On the other hand, sepsis was found to be the sole significant predictor of in-hospital mortality among the other studied exposures, with patients with sepsis having a more than threefold greater risk of in-hospital mortality than those without sepsis. This is consistent with previous reports in which sepsis is a well-established cause of increased mortality in any care setting, not only because of the complications it can cause, but also because of the severity of the risk factors that are usually associated with it (24-26).

The findings of the study have a substantial contribution to the existing literature and can be applied to comparable health care settings for the following reasons: it examined the effect of AKI in non-critical patients where there is a scarcity of evidence, it has a comparator study group for better inference, and it was conducted in a large referral hospital with the best setup for managing patients with AKI that admits patients from all over the country. However, data on details of the stage and control levels of other studied comorbidities that could potentially confound the finding were not studied.

## CONCLUSION

The study found that AKI is not a significant predictor of in-hospital mortality in non-critical care settings, implying that with early detection and proper management, patients with AKI can have a similar prognosis as those without AKI, unlike the critical setting. Sepsis, on the other hand, was identified to be a significant predictor of in-hospital mortality, and clinicians should pay special attention to it in the non-critical setting. However, further large-scale prospective research is required for better understanding and generalizability to a wider context.

## Data Availability

All relevant data are available upon reasonable request from the corresponding author.

## Declaration

### Ethical considerations

The study was conducted after obtaining ethical clearance from St. Paul’s Hospital Millennium Medical College institutional review board (SPHMMC-IRB). Anonymity of the participants was maintained by use of medical record number in the research report. No other personal identifiers of the patients were used in the research report. Access to the collected information was limited to the investigators and confidentiality was maintained throughout the project.

### Consent for publication

Not applicable

### Availability of data and materials

All relevant data are available upon reasonable request from the corresponding author.

### Authors’ Contribution

NDM and TWL conceived and designed the study. NAM, HSB, AAB, KHY, BAH, FNA, LZG, and BGH contributed to the conception and design of the study. NDM and TWL performed statistical analysis, and drafted the initial manuscript. NAM, HSB, AAB, and BAH contributed to the statistical analysis and interpretation of the findings. KHY, FNA, LZG, and BGH revised the manuscript. All authors approved the final version of the manuscript.

## Acknowledgment

The authors would like to thank all individuals involved in the data collection, supervision, and facilitation of the research work.

